# Open science practices in research published in surgical journals: A cross-sectional study

**DOI:** 10.1101/2023.05.02.23289357

**Authors:** Kavya Pathak, Jayson S. Marwaha, Hao Wei Chen, Harlan M. Krumholz, Jeffrey B. Matthews

**Author notes:** **Corresponding Author:** Jayson S. Marwaha, MD, MSc, Department of Surgery, Beth Israel Deaconess Medical Center, Department of Biomedical Informatics, Harvard Medical School, 110 Francis Street, Suite 2G, Boston, MA 02215. Co-first authors (Kavya Pathak, Jayson S. Marwaha). Co-senior authors (Harlan M. Krumholz, Jeffrey B. Matthews).

## Abstract

Open science practices are research tools used to improve research quality and transparency. These practices have been used by researchers in various medical fields, though the usage of these practices in the surgical research ecosystem has not been quantified. In this work, we studied the use of open science practices in general surgery journals. Eight of the highest-ranked general surgery journals by SJR2 were chosen and their author guidelines were reviewed. From each journal, 30 articles published between January 1, 2019 and August 11, 2021 were randomly chosen and analyzed. Five open science practices were measured (preprint publication prior to peer-reviewed publication, use of Equator guidelines, study protocol preregistration prior to peer-reviewed publication, published peer review, and public accessibility of data, methods, and/or code). Across all 240 articles, 82 (34%) used one or more open science practices. Articles in the *International Journal of Surgery* showed greatest use of open science practices, with a mean of 1.6 open science practices compared to 0.36 across the other journals (p<.001). Adoption of open science practices in surgical research remains low, and further work is needed to increase utilization of these tools.

## Introduction

Reproducibility and transparency have long been considered important considerations in medical research.^1^ Recent retractions of studies in several medical journals have underscored the relevance of these issues.^2^ Many tools now exist to promote research quality and transparency, including protocol preregistration websites, Equator Network reporting guidelines for common study types, and preprint servers.^3^ However, the extent to which surgical research has adopted these tools is unknown. The purpose of this study is to describe the use of these quality-promoting practices in surgical research.

## Methods

The use of five open science practices were measured (preprint publication prior to peer-reviewed publication, use of Equator guidelines, study protocol preregistration prior to peer-reviewed publication, published peer review, and public accessibility of data, methods, and/or code) in surgical journals and manuscripts. The top eight general surgery journals by Scimago Journal Rank (SJR2) were included. A random sample of 240 original research articles published from January 1, 2019 to August 11, 2021 from these journals (30 from each journal) comprised the study cohort. The number of journals and studies that explicitly endorsed or used these practices was measured. The study was reported using STROBE guidelines. It was pre-registered on osf.io.

## Results

In their author guidelines, seven (88%) journals recommended the use of Equator guidelines prior to journal submission. Five (63%) journals explicitly stated that they allowed for submissions to have been previously released as preprints. Seven (88%) of journal recommended that authors preregister protocols for clinical trials, though only two (25%) recommended that authors preregister their protocols for observational studies. None (0%) published peer reviewer comments. Four (50%) explicitly recommended that their authors make their methods publicly available. Across all 240 articles, 82 (34%) used one or more open science practices. 63 (26%) explicitly complied with the appropriate Equator guideline. Only 12 (5%) prospective studies preregistered their study protocols. None of the articles were posted on a preprint server prior to journal publication. Only 22 (9%) studies fully disclosed their methods in the form of making their code public or publishing a separate protocol **(Table 1)**. Research in the *International Journal of Surgery* exhibited the highest utilization of open science practices. Studies in this journal utilized a mean of 1.6 open science practices compared to 0.36 across the other journals (p<.001). Journals that recommended or required usage of Equator guidelines saw higher levels of open science practice utilization in their research than journals that did not mention the practice (30% vs. 0% utilization, p value <.001). There was no relationship between journal rank and utilization of open science practices in each journal’s published studies.

**Table 1.** Adoption of various open science tools currently used to promote research quality, transparency, and reproducibility.

| Open Science Tool | Journals (n=8)<sup>1</sup> | Original Research Articles (n=240) |
| --- | --- | --- |
| Equator Network guidelines | 7 (88%) | 63 (26%) |
| Preprints<sup>2</sup> | 5 (63%) | 0 (0%) |
| Preregistration of clinical trials | 7 (88%) | 12 (5%) |
| Preregistration of observational studies | 2 (25%) | 26 (11%) |
| Published peer review | 0 (0%) | 0 (0%) |
| Open source methods | 4 (50%) | 22 (9%) |

<sup>1</sup>The eight journals included in our analysis were: *JAMA Surgery*, *Annals of Surgery*, *British Journal of Surgery*, *Journal of the American College of Surgeons*, *Surgery*, *International Journal of Surgery*, *World Journal of Surgery*, and *American Journal of Surgery*.
<sup>2</sup>Adoption of preprint servers was counted among journals if their author guidelines explicitly allowed for submission of preprint papers, and use of preprint servers was counted among original research articles if it had been posted on a preprint server prior to peer-reviewed publication.

## Discussion

Surgical research is adapting slowly to open science practices in academia, leaving the field potentially vulnerable to poor research quality. We found that surgical journal guidelines vary in their recommendations of open science practices, and use of these practices remains low. Drivers of this lack of open science usage may include limited incentives to follow open science practices, as well as added time required to do so.^4^ To increase usage, researchers, journals, and policymakers can collaborate to provide incentives for open science use, such as publicly available badges, and can advocate for funding structures that require open science usage.^5,6^

This study has limitations, as we only reviewed a sample of published research and excluded subspecialty journals, which may have led us to underestimate open science usage. Despite this, this study demonstrates a lack of open science usage in general surgery journals. The responsibility falls on both researchers and journals to adopt these new tools to promote high quality research generation and dissemination in surgery.

## Data Availability

All data produced are available online at

https://github.com/kavyapathak288/open-sci-surgery

## Supplemental Methods

### Cohort selection

We selected eight general surgery journals with the highest Scimago Journal Rankings (SJR2) in the subject area “surgery” as ranked in 2021.^1^ Affiliate open-source publications were excluded (e.g. British Journal of Surgery Open was excluded). We chose a random sample of 240 articles (30 from each journal) out of all articles published in every issue of these journals from January 1, 2019 to August 11, 2021, to form the study cohort.

### Data collection

We focused on five open science practices: 1) preprint publication prior to peer-reviewed publication, 2) use of Equator network guidelines, 3) study protocol preregistration prior to peer-reviewed publication, 4) published peer review, and 5) public accessibility of data, experimental methods, and/or code.^2^ One authors (KP) reviewed guidelines for authors for each of the eight journals and independently determined whether each set of guidelines did not mention, recommended, or required these open science practices. Journal guidelines that explicitly allowed preprint publication were classified as “recommending” the practice.

For each article in the cohort, two authors (HWC, KP) independently reviewed each study and evaluated whether the same open science practices were followed. Articles were classified as having a published preprint if such a study was cited in the background or methods, or if the study was posted on a preprint server prior to peer-reviewed publication. Studies were classified as using Equator network guidelines if the appropriate guideline for the study (such as PRISMA, CONSORT or STROBE) was mentioned in the methods section. Randomized controlled trials were classified as having a preregistered study protocol if the date the study was posted in clinicaltrails.gov (or an equivalent international registry) was prior to the listed study start date. Observational studies were classified as having a preregistered study protocol if such a protocol was mentioned in the manuscript or posted on a protocol database such as protocols.io, PROSPERO, researchregistry.com, or osf.io. Studies were classified as having published peer review if this was mentioned in the manuscript. Public accessibility of data, methods, or code was assessed by evaluating supplemental materials. Articles were classified as meeting this practice if raw data, code, or detailed study methods were published along with the article, or if it was indicated that data would be made available on request.

Any scoring differences in article classification were reconciled by a third author who independently reviewed the article (JSM). Average inter-rater percentage agreement across all five categories was 97.3%, with 15% of scored items requiring reconciliation. Comparison testing between journals was conducted by ANOVA. Comparison testing between groups of journals (journals that recommended or required compliance with EQUATOR guidelines versus those that did not mention EQUATOR guidelines) was conducted using a *t*-test. All tests used a significance threshold of 0.05. The relationship between SJR2 and utilization of open science practices was determined through linear regression. Inter-rater agreement was calculated using R (version 4.2.2), and all other statistical analyses were conducted in Microsoft Excel (version 2301).

### Methodologic limitations

In this study, we used a random sample of published articles across eight surgical journals, and therefore we did not evaluate all studies published in surgical literature. We did not include subspecialty journals or other general surgery journals. Our study is also limited by whether authors mentioned the use of Equator guidelines, preprints, or preregistration in the text of the article. It is possible that authors used Equator guidelines or submitted preprints but did not mention this in the final manuscript. While we searched preregistration databases such as protocols.io, PROSPERO, and researchregistry.com, it is possible that studies were preregistered in other locations.

### Data availability

The full dataset for this study has been made publicly available and can be viewed and downloaded here: https://github.com/kavyapathak288/open-sci-surgery.

